# Frameworks for measuring population health: a scoping review

**DOI:** 10.1101/2022.11.17.22282470

**Authors:** Sze Ling Chan, Clement Zhong Hao Ho, Nang Ei Ei Khaing, Ezra Ho, Candelyn Pong, Calida Chua, Zongbin Li, Trudi Lim, Sean Shao Wei Lam, Lian Leng Low, Choon How How

## Abstract

**Introduction:** Many regions in the world are using the population health approach and require a means to measure the health of their population of interest. Population health frameworks provide a theoretical grounding for conceptualization of population health and therefore a logical basis for selection of indicators. The aim of this scoping review was to provide an overview and summary of the characteristics of existing population health frameworks that have been used to conceptualize the measurement of population health.

**Methods:** We used the Population, Concept and Context (PCC) framework to define eligibility criteria of frameworks. We were interested in frameworks applicable for general populations, that contained components of measurement of health with or without its antecedents and applied at the population level or used a population health approach. Eligible reports of eligible frameworks should include at least domains and subdomains, purpose, or indicators. We searched 5 databases (Pubmed, EMBASE, Web of Science, NYAM Grey Literature Report, and OpenGrey), governmental and organizational sites on Google and websites of selected organizations using keywords from the PCC framework. Characteristics of the frameworks were summarized descriptively and narratively.

**Results:** Forty-eight frameworks were included. The majority originated from the US (42%), Canada (23%) and Europe (23%). Apart from 1 framework developed for rural populations and 2 for indigenous populations, the rest were for general urban populations. The numbers of domains, subdomains and indicators were highly variable. Health status and social determinants of health were the most common domains across all frameworks. Different frameworks had different priorities and therefore focus on different domains.

**Conclusion:** Key domains common across frameworks other than health status were social determinants of health, health behaviours and healthcare system performance. The results in this review serve as a useful resource for governments and healthcare organizations for informing their population health measurement efforts.

## Introduction

Since its emergence in the 1990s, population health has become an increasingly prominent concept in public health discourse, governance, and research. In their seminal paper, Kindig and Stoddart defines population health as an approach to understanding health that transcends the individual, focusing on interrelated factors and conditions shaping the health of a population, including the environment, social and cultural forces, and lifestyle choices (1). In other words, health cannot be fully understood without a contextualisation of socioeconomic and other factors that are shaped by environments and communities (2). This paradigm shift originated during the 1970s-80s and emerged in response to the growing body of evidence on social determinants of health, and shifting social attitudes concerned with social justice and equity (3). In contrast to the traditional biomedical model that focused on individual risk factors of diseases, a population health approach adopts an upstream and preventive approach to achieving health outcomes.

Population health indicators provide a means for governments and organisations to monitor public health, evaluate interventions, and guide population health policies. Summary measures such as life-expectancy are commonly used to measure the health of a population and for benchmarking against others but are limited on their own (4). With health and its antecedents being complex and multifaceted constructs, the selection of relevant population health indicators is not straightforward. In a scoping review of population health indices, only 7 out of 27 indices had a theoretical or conceptual foundation guiding the aggregation of indicators in a meaningful way (5).

A framework should therefore precede indicator selection (4). Frameworks provide a structure by which to organise the dynamic and interrelated factors between individuals and their environment, and through which to develop hypotheses about how such relationships affect health outcomes over time (6). For instance, the widely accepted Canadian Institutes of Health Research population health framework provides an integrated view of health through upstream forces, proximal causes of heath, life course processes, disparities across sub-populations, health services, and health outcomes, as well as the indicators and indices used to measure them (7). Others may differ depending on their purpose and definition of health and population health.

The usage of a population health framework is necessary as it provides a theoretical grounding and context for selection of indicators and clarifies the role of each indicator (5). Indeed, this is a step many governments and organizations have taken in their population health efforts. There have been reviews on population health indicators (5,7,8). However, to our knowledge there is no work that organises and clarifies this growing body of literature.

In this paper, we conducted a scoping review with the aim of providing an overview and summary of the characteristics of existing population health frameworks that have been used to conceptualize the measurement of population health. Specific aims included to understand what domains are included in the frameworks, how or why they were chosen, and what some representative indicators under each domain were.

## Methods

This scoping review follows the guidelines described by the Preferred Reporting Items for Systematic reviews and Meta-Analyses extension for Scoping Reviews (PRISMA-ScR) checklist (9).

### Eligibility criteria

The eligibility criteria of *population health measurement frameworks* were guided by the elements of the Population, Concept, and Context (PCC) framework. In the population element, we were interested in frameworks that were applied to general populations, which included subsets by demographic variables (e.g. age or ethnicity). However, we excluded populations which were defined by illnesses or diseases (e.g. stroke or mental health patients), or settings (e.g. workplace, schools).

For the Concept element, frameworks should contain components of measurement of health, with or without its antecedents. Frameworks by definition convey structure, at least in the form of categorization (6). Therefore, eligible frameworks should fulfil this definition. Simple lists of indicators without categories are excluded. Frameworks should also be novel, so mere representations of known literature or frameworks with insufficient explanation, and logic models for specific programs were excluded. For context, frameworks should be applied at the macrolevel, or use a population health approach.

Eligible *reports* of eligible frameworks would need to include at least one of the following dimensions – 1) Domains and subdomains; 2) purpose of the framework; or 3) population health indicators used. Where there were more than 1 report for the same framework, we selected the one with the most relevant and comprehensive information. If another report supplemented information not found in this primary report, we would include both. We included primary articles of any study design, reviews and selected grey literature. Conference abstracts, theses and dissertations, letters to editors, commentaries, non-English articles, and articles published before 1990 were excluded.

### Information sources

We searched MEDLINE (PubMed), EMBASE, Web of Science, NYAM Grey Literature Report and OpenGrey databases. In addition, we searched governmental and organizational sites on Google (site:.gov OR site:.org OR site:.net OR site:.eu) and websites of the following governments and organizations known to have population health initiatives and/or frameworks:

- UK National Health Service (NHS)
- Agency for Healthcare Research and Quality (AHRQ)
- Centres for Disease Control (CDC)
- US Department of Health and Human Services
- Public Health Agency of Canada
- Australian Government Department of Health
- World Health Organization (WHO)
- Organisation for Economic Co-operation and Development (OECD)
- Public Health England
- European Union (EU) CDC
- National Quality Forum (NQF)
- Health Information Technology, Evaluation, and Quality Center (HITEQ)

### Search strategy

We used the keywords ‘framework’ and ‘population health’ from the concept and context elements as search terms, respectively. Depending on the database, we used these terms as keywords or also included controlled vocabulary that corresponded to them. The keywords or controlled vocabulary were combined using the BOOLEAN operator ‘OR’ and ‘AND’ within and across the PCC elements, respectively. Where possible, filters were applied to select only human studies, English articles and articles published after 1 Jan 1990. The final search of the databases was performed on 01 December 2021. For some databases (Pubmed, EMBASE, Web of Science) we further applied a ‘title/abstract’ filter to improve the specificity of the search results. If we came across reports that mention an eligible framework but did not contain the relevant details to be included, we then searched for reports on that particular framework. We also searched reference lists of included reports.

### Selection of sources of evidence

Three reviewers (SLC, CZHH, NEEK) developed and piloted the search strategy. Two stages of screenings were performed to select the sources of evidence. At the first stage, the titles and abstracts of each source was screened and selected for full text review by two reviewers independently. In the second stage, the full texts of articles selected in the first stage were also reviewed by 2 reviewers independently. In both stages, a third reviewer would make the final decision in the event of a conflict.

### Data charting process

A data charting form to extract data of interest was developed by one reviewer (SLC) and piloted by another (CZHH). Data from each report was extracted by one reviewer and reviewed by a second reviewer. Any discrepancies were resolved by consensus between the data extractor and reviewer.

### Data items

The data items included citation details, details on the framework (e.g. name, country of origin, organization that developed it, type of population it is applicable to, purpose, approach to development, dimensions in framework apart from domains), and the domains and indicators used in the framework, including definitions or descriptions where available. For domains, we recorded up to 2 further levels of sub-domains (total 3 levels).

### Synthesis of results

To facilitate summary and presentation of results, some variables were reduced to a smaller number of categories manually by a single reviewer (SLC). These variables were the type of organization developing the frameworks, types of population the framework was applicable to, purpose and dimensions of the framework. Types of organizations were broadly categorized into governmental, academic, non-government organizations, non-profit organizations, intergovernmental organizations, and private foundations. Populations were grouped in to general, rural and indigenous populations. While all population health frameworks are tools for measuring, understanding and then improving population health, they were categorized into one of these three main purposes based on the stated purpose.

Finally, dimensions cut across domains and indicators and we focused mainly on a lifespan and equity approach. For the lifespan approach, this generally involve diving into indicators relevant for different life stages and/or breaking down indicators by age groups. For the equity approach this typically involves examining indicators by certain socioeconomic factors. Other specific dimensions mentioned were also included.

Domains and subdomains were also aggregated by concept manually for purposes of visualization. The characteristics of the frameworks were then summarized descriptively using counts and proportions, and median and ranges, as appropriate. The dominant domains and number of domains, subdomains and indicators were visualized using a word cloud and heatmap, respectively. Other aspects of the frameworks were summarized narratively.

## Results

### Search results

A total of 48 population health measurement frameworks were included in this review (Figure 1). The details of the frameworks are shown in Table 1. The full list of the domains, subdomains and indicators are provided in supplementary file 1.

**Figure 1.**
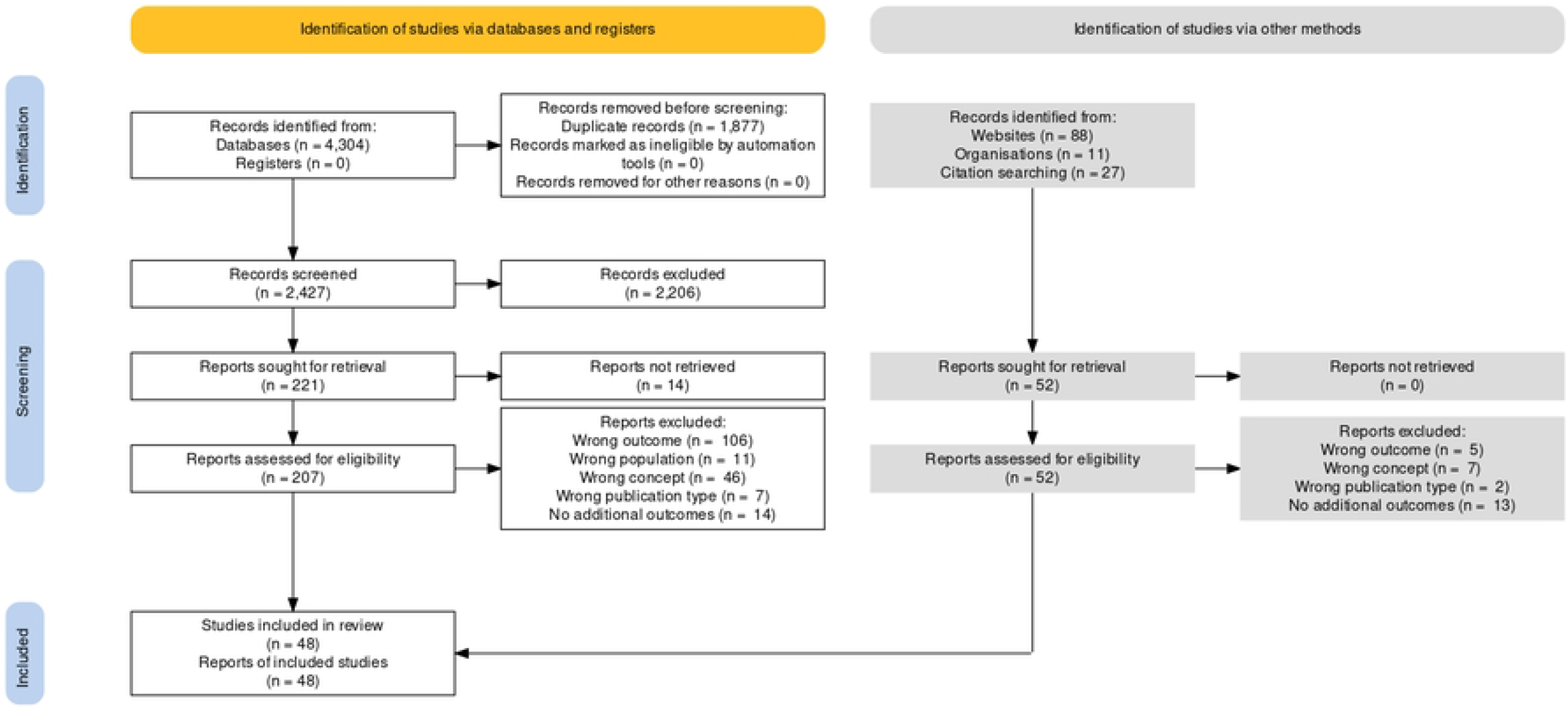
PRISMA diagram

**Table 1.**
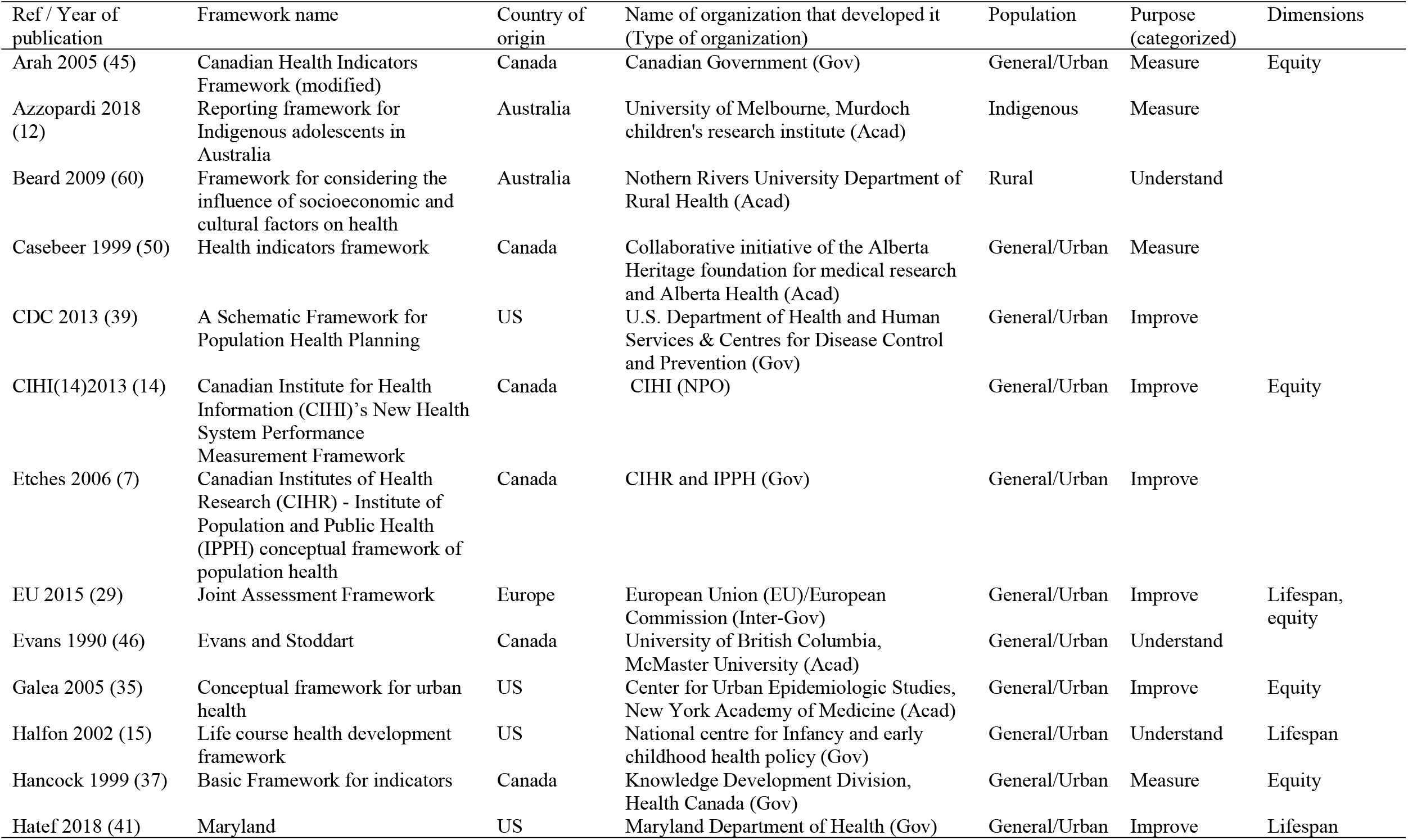
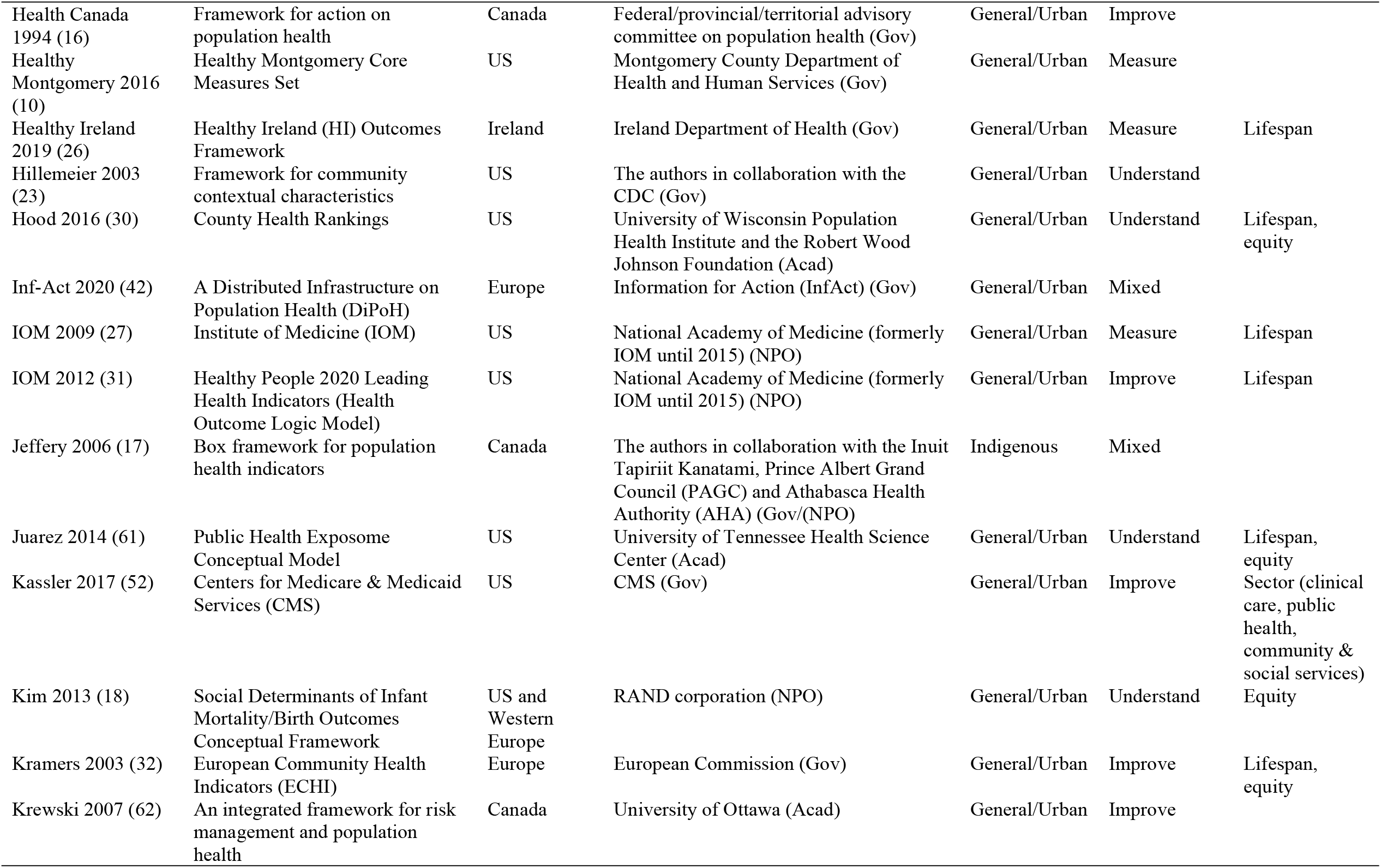
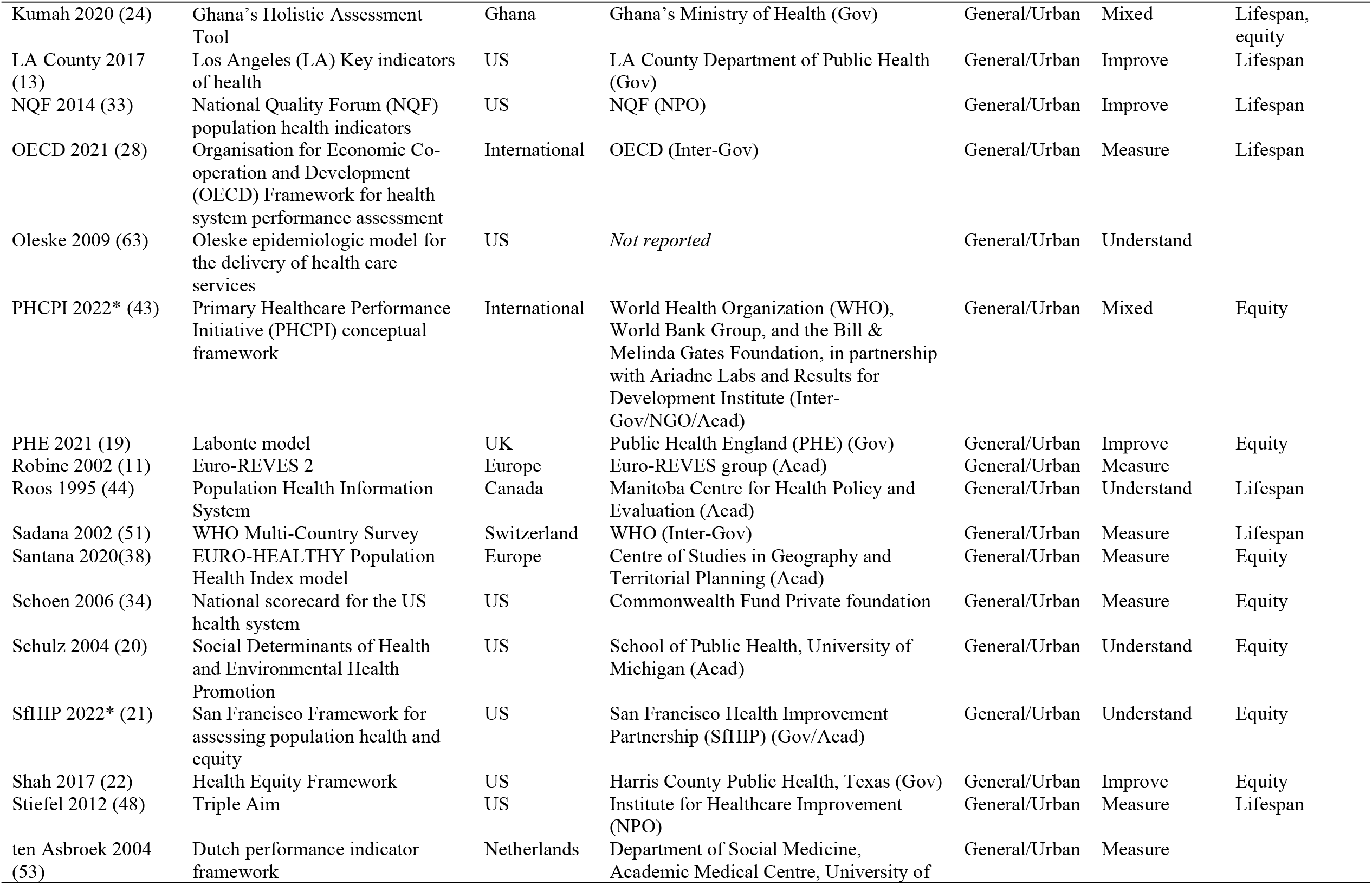
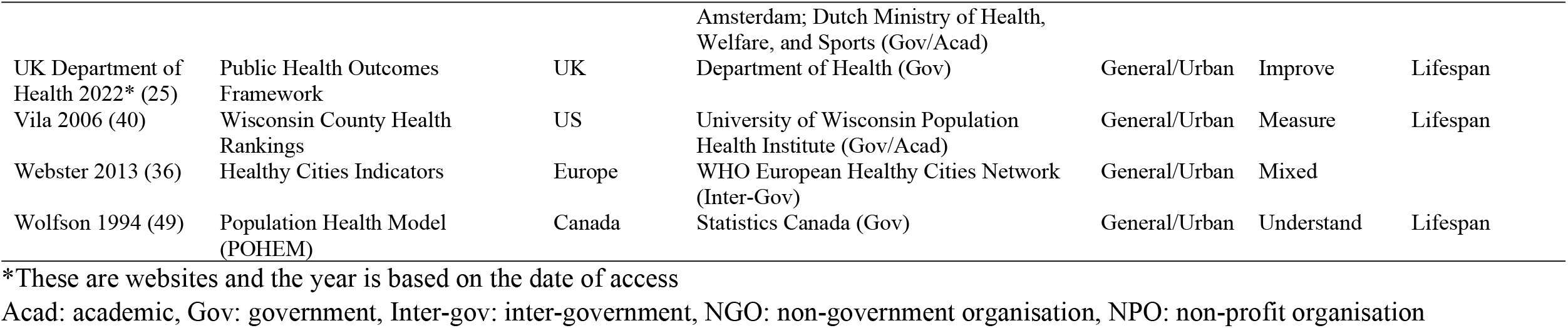
Details of included frameworks

### Characteristics of population health measurement frameworks

Table 2 shows the descriptive statistics of the key characteristics of the frameworks. Majority of the frameworks originated from the US (41.7%), Canada (22.9%) and Europe (22.9%). None were from Asia. Most were published between 2001 and 2020 (77.1%). Governmental (including intergovernmental) and academic organizations accounted for majority of framework development (85.4%). Only three frameworks were developed for specific populations (2 for indigenous and 1 for rural), while the rest were for the general or urban population. In terms of purpose, the frameworks were fairly evenly distributed across the three categories. Two-thirds of the frameworks mentioned some dimension, and these were also distributed quite evenly between lifespan and equity approaches.

**Table 2.** Summary of key characteristics of frameworks

| Characteristics | N (%) |
| --- | --- |
| Year of publication |  |
| 2000 and before | 6 (12.5) |
| 2001 to 2010 | 17 (35.4) |
| 2011 to 2020 | 20 (41.7) |
| 2020 onwards | 5 (10.4) |
| Country of origin |  |
| US | 20 (41.7) |
| Canada | 11 (22.9) |
| Europe | 11 (22.9) |
| International | 2 (4.2) |
| Australia | 2 (4.2) |
| US and Western Europe | 1 (2.1) |
| Ghana | 1 (2.1) |
| Type of organization framework originated from |  |
| Governmental | 19 (39.6) |
| Academic | 12 (25.0) |
| Non-profit organization | 6 (12.5) |
| Intergovernmental | 4 (8.3) |
| Governmental/academic | 3 (6.3) |
| Governmental/non-profit organisation | 1 (2.1) |
| Intergovernmental/academic/non-governmental organisation | 1 (2.1) |
| Private foundation | 1 (2.1) |
| Population framework is applied to |  |
| General/urban | 45 (93.8) |
| Indigenous | 2 (4.2) |
| Rural | 1 (2.1) |
| Purpose |  |
| Improve | 16 (33.3) |
| Measure | 15 (31.2) |
| Understand | 12 (25.0) |
| Measure/improve | 2 (4.2) |
| Measure/understand | 2 (4.2) |
| Understand/improve | 1 (2.1) |
| Dimensions |  |
| None | 16 (33.3) |
| Lifespan | 14 (29.2) |
| Equity | 12 (25.0) |
| Lifespan and equity | 5 (10.4) |
| Sector | 1 (2.1) |

### Domains and subdomains

Figure 2 shows the numbers of domains, sub-domains and indicators for each framework. Majority of the frameworks have between 1 to 5 domains (68.8%) but have more level 2 sub-domains (31.2% have 6-10, 27.1% have 11-20 and 18.8% have >20). The median number of domains and level 2 subdomains are 4 (range 2 – 14) and 9.5 (range 0 -65), respectively. Half of the frameworks do not have level 3 subdomains. Of those that do, most have >10 (75%). The median number of indicators is 19.5 (range 0 – 255). Twenty-one of the frameworks did not have indicators (43.8%). Of those that do, majority have >20 indicators (85.2%).

**Figure 2.**
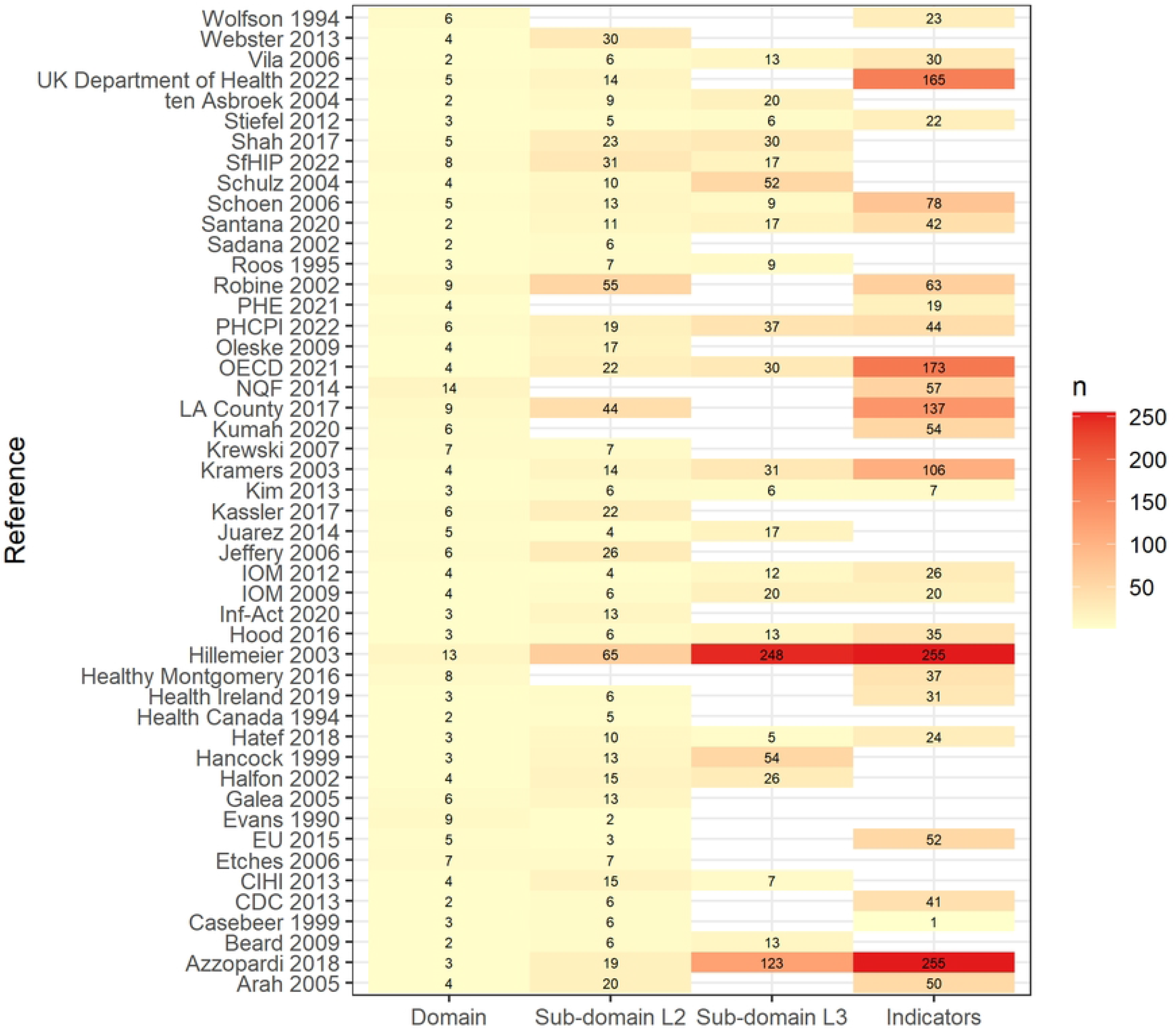
Heatmap of number of domains, subdomains and indicators L2: level 2, L3: level 3.

Figure 3 shows the word cloud for the domains. The most common concepts were health, context or social determinants of health, health behaviours, and those related to the healthcare system. For health, most frameworks used summary indicators of health such as mortality and life-expectancy, and indicators of a few selected health conditions. However, four frameworks had longer lists of indicators for specific communicable and non-communicable diseases (10–13). Of note, psychological or mental health risk factors and/or outcomes feature in 25 (52%) of the frameworks, highlighting its emerging importance (10– 34).

**Figure 3.**
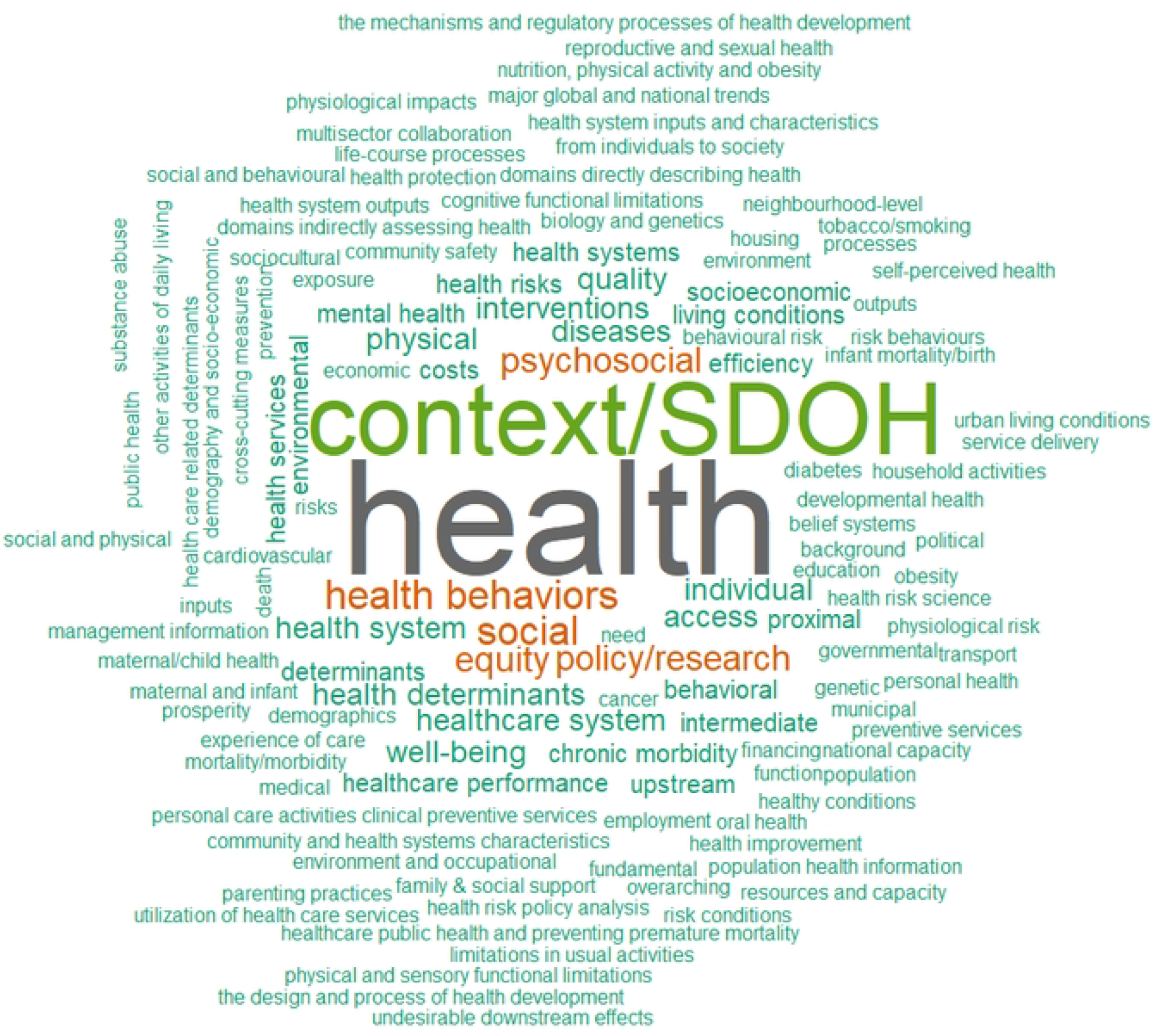
Wordcloud for framework domains SDOH: social determinants of health.

The social determinants of health domain were present under some label or other in all except the Ghana Holistic Assessment Tool and Euro-REVES 2 framework (11,24). Some of the frameworks elaborate on these factors, with sub-domains and indicators on the physical environment, social environment, and even politics, national and global trends (17,20–23,35– 38). For example, the conceptual framework for urban health measures sub-domains such as immigration, globalization and the changing role of government (35). The framework for community contextual characteristics, one of the two frameworks with the largest number of indicators, also measures the economic, employment, education, political, environmental, housing, governmental, transport aspects in the region where the population of interest is located (23). Interestingly, crime features in 10 frameworks, as this affects the physical safety of people in a community (12,13,20,23,25,30,33,38–40). Many frameworks also measure lifestyle and health-related behaviours. Apart from the common ones like diet, physical activity, smoking and alcohol use, some frameworks include sexual behaviour, use of illicit drugs, seatbelt behaviour, immunization or health screening, breastfeeding and induced abortion (10,12,21–23,25,26,30–33,39). One even included measures of parenting practices (13).

A quarter of the frameworks have domains that pertain to the healthcare system or healthcare performance. One example is the OECD framework, which assesses health system performance within the context of other contextual determinants of health (28). Within the construct of healthcare performance, common subdomains are accessibility, capacity, quality, cost and effectiveness (13,27–29,32,34,38,41–45).

A few of the frameworks had specific focuses and therefore unique domains and indicators that are relevant largely for their setting. For example, the reporting framework for indigenous adolescents in Australia contained domains that were largely relevant for that community, such as ‘family, kinship and community health’, which explored family roles and responsibilities, contact with extended family, removal from family, participation in community events and sense of belonging to the community (12). Another example is the Ghana’s Holistic Assessment Tool, which contains indicators for health-related United Nations sustainable development goals (SDGs) such as proportion of deliveries attended by a trained health worker, proportion of children under 5 years sleeping under insecticide treated net, and tuberculosis treatment success rate, and certain endemic communicable diseases such as non-acute flaccid paralysis polio rate (24).

### Approach to framework development

Evans and Stoddart developed the first population health measurement framework in 1990 (46) based on a much earlier 1974 Whitepaper titled “A new perspective on the health of Canadians”, which recognized the limitations of the healthcare system on improving health status and presented a preliminary framework of the ‘health field’ (47). Subsequent frameworks were mostly developed from one or a combination of four approaches: 1) adaptation from an existing framework (11,12,19–22,28,31,33,40,43,44,48,49), 2) environmental scan of existing frameworks and literature review to summarize current knowledge of health determinants (7,14,16,27,29,40,41,43,50–53), 3) consulting and getting inputs from experts and stakeholders (12,14,23,25,26,29,32,34,38,40,41,43,51) and 4) basing on past work, priorities and goals of the organization developing it (7,18,35,36,45,53).

## Discussion

Population health has been a popular concept in healthcare for the past 3 decades but interestingly does not have a unanimous definition (1,2,54). The most commonly used definition, which originated from Kindig and Stoddart, defines population health as ‘the health outcomes of a group of individuals, including the distribution of such outcomes within the group” (1). Nevertheless, people working on ‘population health’ would have different focuses, goals and populations of interest (54). This may explain the large number of population health frameworks we found in this review.

The concept of population health purportedly originated from Canada, so it was unsurprising majority of the frameworks came from Canada and US. Many of these efforts were also motivated by the articulation of the Triple Aims as a goal for the US healthcare system in the late 2000s, in which improving population health was one of them (55).

Health status and social determinants of health were the most common domains across the frameworks. As seen from the word cloud, there were also many other domains that were closely related to and/or could be considered subdomains of one of these domains. This is because different frameworks have different level of detail, and the hierarchy of domains and subdomains are different in level of detail across frameworks. In other words, a subdomain in one framework could be a domain in another, or an indicator in one framework could be a subdomain in another. It is therefore also difficult to summarize domains and subdomains in a simple way across the frameworks.

The domains and subdomains chosen in different frameworks therefore largely reflects the current knowledge of population health, what influences it, and the focus of the organization(s) developing it. It is unsurprising to see that some key domains appear in many frameworks while different domains expanded to varying detail. For example, social determinants of health features in all frameworks except two frameworks. Some frameworks have a heavy focus on health status, such as the Healthy Montogomery Core Measures Set, Triple Aim and Euro-REVES 2, with the Euro-REVES 2 framework even measuring activities of daily living and degree of functional limitations (10,11,48). Other frameworks break down the social determinants into considerable detail, such as the framework for community contextual characteristics, life course health development framework, Healthy Cities Indicators, and others (15,18–20,22,23,36–38,40). Several have a heavier focus on healthcare performance, such as the EU Joint Assessment Framework, European Community Health Indicators (ECHI), OECD, the Primary Healthcare Performance Initiative (PHCPI), National scorecard for the US health system (28,29,32,34,43). Others are generally more balanced between the domains.

The results of this scoping review can serve as an evidence base for governments and/or health systems developing their own population health measurement frameworks and selecting indicators for their population health initiatives. They can select and adapt from the frameworks available, and assess the relevance of the range of domains, subdomains and indicators in their context. Populations are largely unique as they are shaped by their local and wider contextual factors. As such, no one framework used in one population or healthcare system is likely directly applicable to another population or healthcare system without adaptation. Population health practitioners can derive any level of detail that matches their interests and requirements from this review, from a broad sense of the literature down to specific indicators.

Settings which are further ahead in the population health journey with existing indicators can also use these results to assess what domains and subdomains have been covered, and where the gaps are. For example, population health is an increasingly important national priority in Singapore and the Ministry of Health is planning several major initiatives to improve the health of the general population (56,57). To achieve this, the Ministry is working closely with the three major public healthcare clusters in Singapore to develop a set of population health indicators and the evidence base here can help inform the choices. With an initial set of indicators, practitioners can also interrogate their data systems and medical records to determine if they are available or if they need to build prospective data collection tools. This can also be an iterative process for selecting indicators using the results here as a resource. One constraint of the data in its current form though is the difficulty in navigating the long list of domains, subdomains and indicators. In future work, we aim to design a dashboard that allows for interactive exploration of the scoping review data.

There are limitations to this scoping review. Firstly, some frameworks might have been missed due to our language restriction, especially those in Asia. However, many official documents from this region are available in English, so this might not have impacted the search results significantly. Secondly, there are many terms and concepts in the literature that have overlaps with population health, such as public health, urban health, global health, population health management, health equity, health system performance and social determinants of health. Based on our inclusion criteria, concepts like urban health, rural health, community health and global health would be included as they pertain to general populations albeit in different types of settings. Related concepts such as health equity, social determinants of health and health system performance were not the focus of the search and could be part of the frameworks included. However, if a framework was focused on one of these concepts alone without the measurement of health status, then it would be excluded. Some frameworks also focused more on population health management and if it looked more like a logic model for specific interventions then these would also be excluded (58,59). Overall, this review represents a useful collection of frameworks used for measuring the health of a population and its key antecedents. (60)

## Conclusion

We found 48 frameworks for the measurement of population health with variable numbers of domains, subdomains and indicators, and depth of detail. The key domains apart from health status were social determinants of health, health behaviours and healthcare system performance. These results serve as a useful resource for governments and healthcare organizations for informing their population health measurement efforts.

## Data Availability

All relevant data are within the manuscript and its Supporting Information files.

## Acknowledgements

We would like to thank Ms Sabrina Liau for her assistance with article screening.

